# Maternal and neonatal outcomes of pregnant women with COVID-19 pneumonia: a case-control study

**DOI:** 10.1101/2020.03.10.20033605

**Authors:** Na Li, Lefei Han, Min Peng, Yuxia Lv, Yin Ouyang, Kui Liu, Linli Yue, Qiannan Li, Guoqiang Sun, Lin Chen, Lin Yang

## Abstract

**Background:** The ongoing epidemics of coronavirus disease 2019 (COVID-19) have caused serious concerns about its potential adverse effects on pregnancy. There are limited data on maternal and neonatal outcomes of pregnant women with COVID-19 pneumonia.

**Methods:** We conducted a case-control study to compare clinical characteristics, maternal and neonatal outcomes of pregnant women with and without COVID-19 pneumonia.

**Results:** During January 24 to February 29, 2020, there were sixteen pregnant women with confirmed COVID-19 pneumonia and eighteen suspected cases who were admitted to labor in the third trimester. Two had vaginal delivery and the rest took cesarean section. Few patients presented respiratory symptoms (fever and cough) on admission, but most had typical chest CT images of COVID-19 pneumonia. Compared to the controls, COVID-19 pneumonia patients had lower counts of white blood cells (WBC), neutrophils, C-reactive protein (CRP), and alanine aminotransferase (ALT) on admission. Increased levels of WBC, neutrophils, eosinophils, and CRP were found in postpartum blood tests of pneumonia patients. There were three (18.8%) and two (10.5%) of the mothers with confirmed or suspected COVID-19 pneumonia had preterm delivery due to maternal complications, which were significantly higher than the control group. None experienced respiratory failure during hospital stay. COVID-19 infection was not found in the newborns and none developed severe neonatal complications.

**Conclusion:** Severe maternal and neonatal complications were not observed in pregnant women with COVID-19 pneumonia who had vaginal delivery or caesarean section. Mild respiratory symptoms of pregnant women with COVID-19 pneumonia highlight the need of effective screening on admission.

## Introduction

In December 2019, an outbreak of the 2019 coronavirus disease (COVID-19) associated pneumonia was reported in Wuhan, a mega city with an 11 million population in central China, and soon spread to other cities in China and overseas[1]. The causative pathogen was identified as a novel coronavirus, severe acute respiratory syndrome coronavirus 2 (SARS-CoV-2) [1]. As of 06 March 2020, COVID-19 has caused 80,735 confirmed cases in China, of whom 3,045 died and 5,737 were admitted into ICU [2]. In response to this fast-spreading epidemic, the Chinese government has locked down the epicenter Wuhan city since 23 January 2020, and implemented a series of social distancing measures such as strict traffic restrictions, forbidden social gatherings, closure of residential communities[3]. The epidemiological data showed that most cases had mild symptoms, and overall case fatality rate was 2.3%. Although the virulence of SARS-CoV-2 seems considerably lower than two previous zoonotic coronaviruses, SARS-CoV and MERS-CoV, it was far more efficient to transmit between close contacts.[4] Particularly, this novel coronavirus has caused special concerns in pregnant women, because both SARS-CoV and MERS-CoV have been found to cause severe complications in pregnant women[5]. Several reports on suspicious vertical transmission of this virus have further increased such concerns[6]. Although the recent laboratory studies and clinical reports did not find strong evidence to support a vertical transmission route, the possibility still cannot be completely ruled out [7-9].

Clinical and epidemiological features of COVID-19 infection have been widely reported[10-14]. However, the clinical reports on maternal and neonatal outcomes of pregnant women with SARS-CoV-2 infection remain sparse. An earlier study by Chen *et al* reported nine pregnant women with COVID-19 pneumonia, who took cesarean section in a tertiary hospital of Wuhan [7]. These patients showed clinical symptoms similar to non-pregnant patients with COVID-19 pneumonia. They also claimed that there was no evidence of vertical transmission. To date, none of previous studies have compared maternal and neonatal outcomes of pregnant women with COVID-19 pneumonia to those without pneumonia, to investigate the adverse effects of COVID-19 infection on pregnancy.

## Methods

### Study design and patients

We retrospectively reviewed medical records of pregnant women who were admitted into the Hubei Provincial Maternal and Child Health Center, a tertiary hospital in Wuhan with 1,900 hospital beds, during January 24 – February 29, 2020. We followed the clinical diagnosis criteria for COVID-19 pneumonia in the New Coronavirus Pneumonia Prevention and Control Program (5th edition) by the National Health Commission of China[15]. Throat swabs were collected from all these patients and sent to the laboratory of the Wuhan Center for Disease Control and Prevention for tests of SARS-CoV-2. Diagnosis criteria of COVID-19 infection include 1) typical chest CT imaging of patchy shadows and ground glass opacity, and 2) positive in reverse transcription polymerase chain reaction (RT-PCR) tests for SARS-CoV-2. However, previous studies argued that false negative cases might be common for COVID-19 infection cases due to low virus titers, sampling at late stage of illness, and inappropriate swabbing sites[7]. Given overloaded healthcare systems and limited test capacities during our study period, we were concerned about underreporting of COVID-19 cases if solely relying on laboratory tests. Therefore, in this study we also included the suspected patients with typical chest CT imaging but negative in RT-PCR tests. Eleven pregnant women who were tested positive for SARS-CoV-2 were classified as laboratory confirmed case group, and eighteen with typical chest CT imaging but tested negative in RT-PCR tests as suspected case group.

The control group of pregnant women without pneumonia during hospital stay were randomly selected from the medical records by an investigator (MP), who was not involved in statistical analysis. Only those aged 25-35 years were selected to match the age range of cases. We selected 121 women who were admitted during the same period (control 2020 group). Considering the potential adverse effects of mental stress caused by city lockdown and severe epidemics, we also included a second control group of 121 women admitted during January 24 – February 11, 2019 (control 2019 group). Blood test results were also retrieved from medical records. Two case groups underwent a blood test every three days but two control groups only took once on admission.

### Data analysis

Clinical characteristics, laboratory test results, maternal and neonatal outcomes were collected from medical records and reviewed independently by two investigators (YXL and YO). Fisher’s exact tests and Mann-Whitney U tests were used to compare the group difference for categorical and continuous variables, respectively. Friedman tests were used to test for the difference of blood test results across time within the same subjects. All the data analysis was conducted using R version 3.6.2.

## Results

### Clinical characteristics and maternal outcomes

Demographic characteristics of two case groups and two control groups are shown in Table 1. The age of confirmed cases ranged 26-37 years and all were in the third trimester. Two confirmed cases (18.2%) and one suspected case had chronic conditions of hypertension, polycystic ovary syndrome and hepatitis B. Their gestational weeks on admission ranged 33 weeks plus 6 days to 40 weeks plus 4 days. Around 70% of two case groups had other maternal complications, significantly higher than the controls (31-33%). All these complications were developed before diagnosis of pneumonia.

**Table 1.** Demographic characteristics of two case groups and two control groups.

|  | Confirmed cases<br>(n=16) | Suspected cases<br>(n=18) | *P* value <sup>a</sup> | Control 2020<br>(n=121) | *P* value <sup>a</sup> | Control 2019<br>(n=121) | *P* value <sup>a</sup> |
| --- | --- | --- | --- | --- | --- | --- | --- |
| Age (years ± SD) | 30.9 ± 3.2 | 29.8 ± 2.3 | 0.624 | 30.1 ± 3.3 | 0.501 | 29.3 ± 2.6 | 0.090 |
| Caesarean delivery (n, %) | 14 (87.5%) | 16 (88.9%) | 1.000 | 57 (47.1%) | 0.003 | 44 (36.4%) | <0.001 |
| Chronic illness (n, %) | 2 (12.5%) | 1 (5.6%) | 0.591 | 5 (4.1%) | 0.190 | 0 (0.0%) | 0.013 |
| Complications in pregnancy (n, %) | 11 (68.5%) | 13 (72.2%) | 1.000 | 38 (31.4 %) | 0.005 | 32 (33.3%) | 0.011 |

<sup>a</sup> *P* value of Fisher's exact tests and Mann-Whitney U tests, the laboratory confirmed cases as reference group.

Fourteen patients had caesarean section, because confirmed or suspected COVID pneumonia has become one indication for caesarean section in our hospital since 24 January 2020. Two patients had vaginal delivery because neither presented any respiratory symptoms when admitted for full-term labor. One of them had fever two days after childbirth and another had CT images of patchy shadows in the right lung on the same day of labor.

In addition to pneumonia, eleven out of 16 confirmed cases had gestational complications on admission, including gestational diabetes mellitus (n=3), premature rupture of membranes (1), gestational hypertension (3), hypothyroidism (2), preeclampsia (1) and sinus tachycardia (1). None of confirmed COVID-19 patients reported an exposure history. Four were admitted with fever for investigation and eight developed fever after childbirth (Table 2). None presented other respiratory symptoms on admission nor during hospital stay. Two of suspected COVID-19 pneumonia patients reported cough, sore throat, dyspnea, diarrhea and vomiting.

**Table 2.** Clinical characteristics of pregnant women with confirmed or suspected COVID-19 pneumonia.

| Characteristic | Confirmed cases (%) | Suspected cases (%) |
| --- | --- | --- |
| Length of stay, median (IQR), d | 9 (5.5, 11) | 6 (5, 9.3) |
| **Symptoms** |  |  |
| Fever on admission | 4 (25.0%) | 1 (5.6%) |
| Fever after childbirth | 8 (50.0%) | 6 (33.3%) |
| Cough | 0 (0.0%) | 1 (5.6%) |
| Sore throat | 0 (0.0%) | 1 (5.6%) |
| Dyspnoea | 0 (0.0%) | 1 (5.6%) |
| **CT imaging** |  |  |
| Pneumonia in one lung | 8 (50.0%) | 10 (55.6%) |
| Pneumonia in two lungs | 7 (43.8%) | 7 (38.9%) |
| **Treatment** |  |  |
| Steroid | 0 (0.0%) | 0 (0.0%) |
| Antibiotics | 16 (100.0%) | 18 (100.0%) |
| Antivirals | 1 (6.3%) | 0 (0.0%) |
| High throughput oxygen | 0 (0.0%) | 0 (0.0%) |
| ECMO | 0 (0.0%) | 0 (0.0%) |
| **Outcome** |  |  |
| Discharge | 7 (43.8%) | 18 (100.0%) |
| Transfer | 8 (50.0%) | 0 (0.0%) |
| In treatment | 1 (6.3%) | 0 (0.0%) |

Abbreviations: ECMO, extracorporeal membrane oxygenation.

### Laboratory investigations and treatment

All patients took chest CT scans. Ten of confirmed cases had typical image of pneumonia in both lungs and seven in single lung. Fifteen out of eighteen suspected patients had both lungs affected. Compared to the controls, two case groups had slightly lower counts of white blood cells (WBC), neutrophils, C-reactive protein (CRP) and alanine aminotransferase (ALT) on admission, although none reached statistical significance and most were marginally beyond the normal range (Table 3). Lymphocytes, eosinophils and aspartate aminotransferase (AST) were comparable between the cases and controls. An increase of WBC, neutrophils, and CRP were observed in the first postpartum blood test of confirmed cases, followed by a decrease in the second postpartum test (Figure 1). But this transient change was not found in suspected cases. Lymphocytes remained at the lower end of normal range in two case groups.

**Table 3.** Comparison of blood test results on admission between two case groups and two control groups.

|  | Reference range | Confirmed cases | Suspected cases |  | Control 2020 |  | Control 2019 |  |
| --- | --- | --- | --- | --- | --- | --- | --- | --- |
|  |  | Mean (SD) | Mean (SD) | *P* value <sup>a</sup> | Mean (SD) | *P* value <sup>a</sup> | Mean (SD) | *P* value <sup>a</sup> |
| WBC, ×10<sup>9</sup>/L | 3.5 - 9.5 | 8.6 ± 1.8 | 11.6 ± 4.5 | 0.023 | 10.3 ± 3.1 | 0.021 | 9.6 ± 2.4 | 0.179 |
| Lymphocytes, ×10<sup>9</sup>/L | 1.1 - 3.2 | 1.5 ± 0.4 | 1.3 ± 0.5 | 0.512 | 1.5 ± 0.5 | 0.622 | 1.5 ± 0.4 | 0.404 |
| Lymphopenia (<1×10<sup>9</sup>/L), % |  | 2 (12.5%) | 5 (27.8%) | 0.405 | 15 (12.6%) | 1.000 | 14 (11.6%) | 1.000 |
| Neutrophils, ×10<sup>9</sup>/L | 1.8 - 6.3 | 6.6 ± 1.8 | 9.3 ± 5.0 | 0.098 | 8.4 ± 2.7 | 0.007 | 7.4 ± 2.2 | 0.161 |
| CRP, mg/L | 0 - 4.0 | 4.8 ± 4.8 | 11.1 ± 12.9 | 0.043 | 23 ± 41.5 | 0.194 | 8.5 ± 15.7 | 0.574 |
| Elevated CRP (>4.0 mg/L), % |  | 5 (31.3%) | 11 (61.1%) | 0.100 | 68 (58.1%) | 0.060 | 57 (47.1%) | 0.290 |
| Eosinophils, ×10<sup>9</sup>/L | 0.02 - 0.52 | 0.04 ± 0.05 | 0.02 ± 0.03 | 0.109 | 0.08 ± 0.2 | 0.008 | 0.07 ± 0.05 | 0.002 |
| ALT, U/L | 0 - 40 | 11.6 ± 5.0 | 20.6 ± 40.4 | 0.679 | 11.2 ± 8.9 | 0.236 | 11.6 ± 13.9 | 0.181 |
| AST, U/L | 0 - 35 | 16.3 ± 5.2 | 27.8 ± 45.5 | 0.569 | 18.3 ± 6.2 | 0.129 | 17.2 ± 9.7 | 0.728 |

Abbreviations: WBC, white blood cells; CRP, C- reactive protein; ALT, alanine aminotransferase; AST, aspartate aminotransferase.
<sup>a</sup> *P* value of Fisher's exact tests and Mann-Whitney U tests, the laboratory confirmed cases were reference group.
<sup>b</sup> Data were missing in 2 patients for WBC, 2 for lymphocytes, 3 for neutrophils, 4 for CRP, 4 for eosinophils, 2 for ALT and 2 for AST.

**Figure 1.**
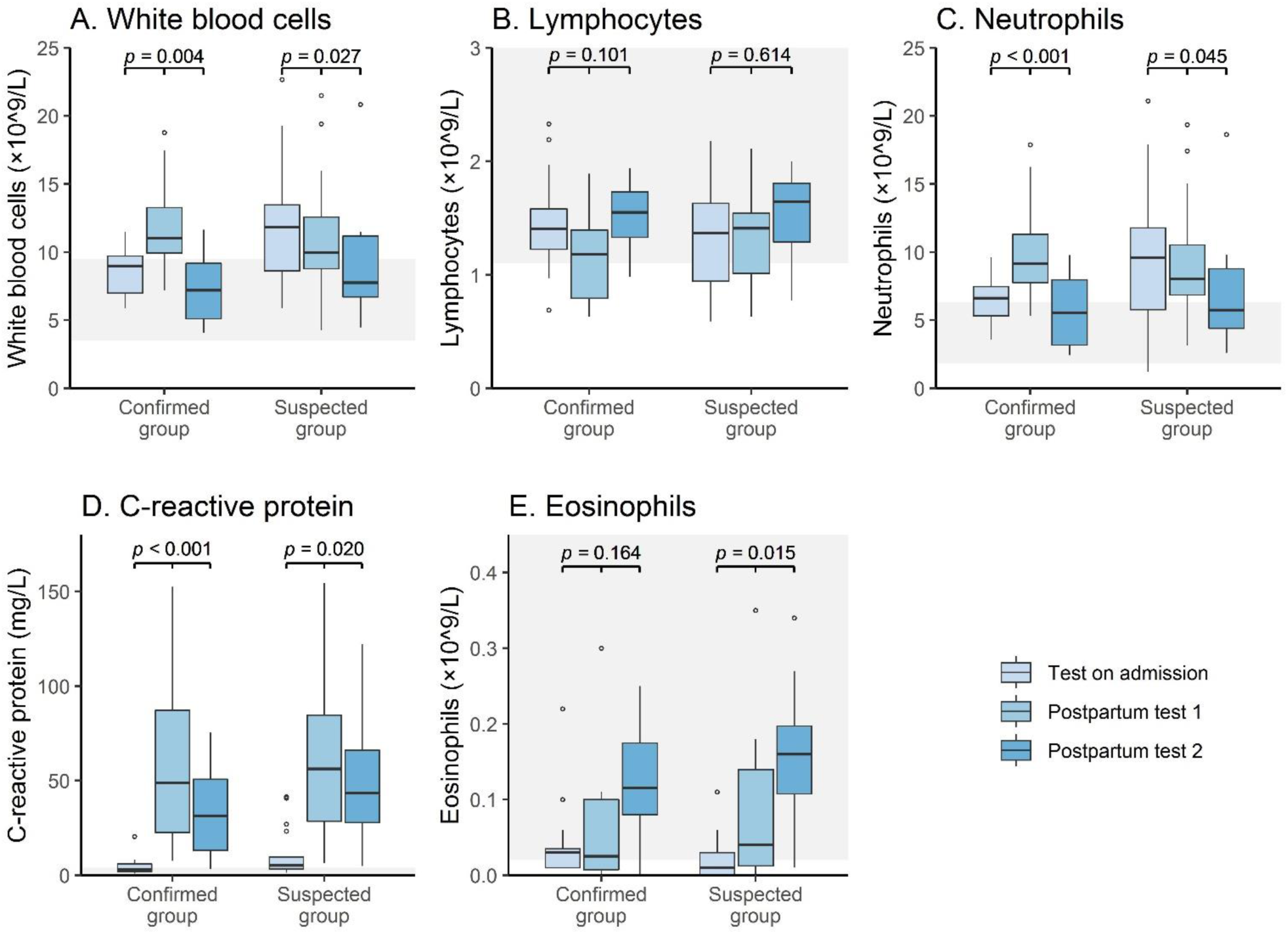
Box plots of blood test results of the confirmed and suspected case groups. Reference ranges of blood tests results are highlighted in grey. *P* values were calculated from Friedman tests for difference of blood tests results across three tests.

All COVID-19 pneumonia patients received antibiotics and four patients received antivirals during hospital stay. All of them have been discharged or transferred to the designated hospitals for COVID-19 patients, and length of stay in our hospital ranged from 3 to 26 days, with a median of 9 days. None were admitted into the intensive care unit (ICU) because of COVID-19 pneumonia or severe maternal complications.

### Neonatal outcomes

Sixteen patients with confirmed COVID-19 pneumonia gave birth to seventeen babies (ten singleton and two twins). Two singletons were born prematurely due to premature rupture of membranes and placental abruption. There were 23.5% and 21.1% preterm births among the newborns of the mothers with confirmed or suspected COVID-19 pneumonia, significantly higher than those of the controls (5.8% and 5.0% in the 2020 and 2019 controls) (Table 4). Low birth weight also occurred more often in two case groups (17.6% and 10.5%) than in two control groups (2.5%). Newborns showed no significant differences between the cases and controls in key neonatal indicators including gestational age at birth, APGAR score at 5 minutes, and intrauterine fetal distress. No events of severe neonatal asphyxia and deaths occurred in these newborns.

**Table 4.**
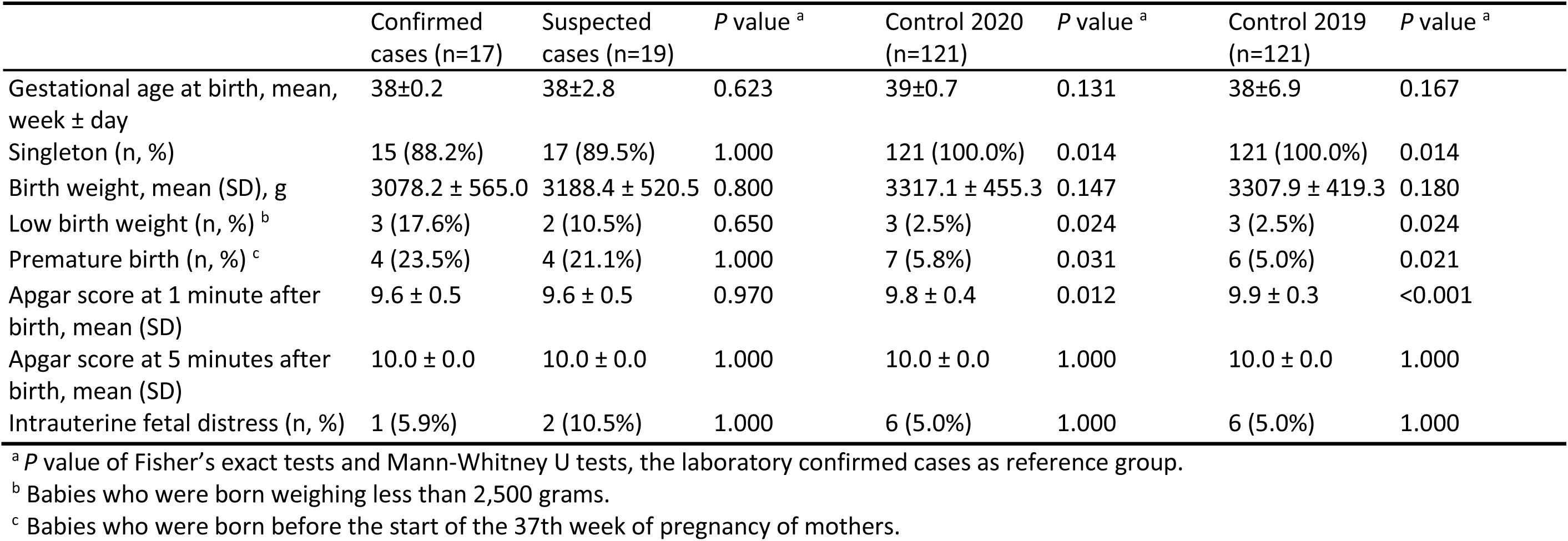
Clinical characteristics of newborns from two case groups and two control groups.

To reduce contact transmission, all COVID-19 patients were immediately moved to isolation wards after delivery or cesarean section, and their newborns were taken care by other family members. Three newborns (including two twins), who were delivered by caesarean section, took throat swabs at 4 and 14 days after birth. All three tested negative for SARS-CoV-2.

## Discussion

To our best knowledge, this is the first case-control study to comprehensively compare maternal and neonatal outcomes of COVID-19 pneumonia with the patients with non-COVID-19 pneumonia and without pneumonia. We found that SARS-CoV-2 infection caused generally mild respiratory symptoms in pregnant women. Clinical signs and symptoms mainly included fever and pneumonia, but other respiratory symptoms were less common. Our results echo the findings of a previous study in pregnant women with SARS in Hong Kong, reporting that fever was the dominant presenting symptom[16]. However, it is of note that most patients did not have any symptoms on admission. For the purpose of screening for suspected cases, we asked all pre-laboring pregnant women to take low-dose chest CT scans with the abdominal region covered, and found that 2.1% fulfilled the criteria of COVID-19 pneumonia (patchy shadows and ground glass opacity in single or both lungs). This highlights the need of enhancing screening for COVID-19 pneumonia on admission, as well as strengthening infection control measures in obstetric wards during the epidemics.

Compared to other COVID-19 pneumonia patients, pregnant women generally had no or mild respiratory symptoms. None of our patients developed severe respiratory complications to require critical care. Laboratory investigations on admission found lower counts of WBC, neutrophils, CRP, and ALT in pregnant women, compared to the non-pneumonia controls. These findings are consistent with those reported in other hospitalized COVID-19 patients who often had lymphopenia and decreased WBC [10]. Slightly increased WBC, neutrophils, eosinophils and CRP were found in postpartum blood tests. We also notice that confirmed and suspected cases shared similar dynamic profiles, suggesting that laboratory test results might not be very useful in making differential diagnosis.

An earlier clinical descriptive study by Chen *et al*. reported the clinical features of nine pregnant women with laboratory confirmed COVID-19 infection cases, all of whom took cesarean section in one tertiary hospital in Wuhan.[7] In our study, in addition to nine cesarean section patients, we also reported two pregnant woman who had a full-term vaginal delivery and was confirmed with COVID-19 pneumonia on the day of delivery. We observed a higher incidence rate of premature delivery in confirmed cases (33.3%), but none was due to severe maternal respiratory failure. This rate was higher than in suspected patients (21.1%) and then two control groups (∼5%) in our study, but slightly lower than the rate of 44% in confirmed COVID-19 pneumonia patients reported by Chen *et al*.[7] We did not observe any deaths or events of severe complications associated with COVID-19 pneumonia that required critical care in the pregnant women and newborns. Hence, the adverse effects of COVID-19 pneumonia on pregnancy appear less severe than those of SARS-CoV and MERS-CoV. There were three pregnant women died during the 2003 SARS outbreak in Hong Kong, and preterm delivery was as high as 80% [17]. Although no maternal deaths were recorded in the MERS-CoV outbreak, more than half of their newborns required critical care and nearly 30% eventually died[18]. A previous study also reported that SARS-CoV infection could increase the risk of preterm delivery in second trimester and spontaneous abortion in first trimester[16]. Since all of our patients were in the third trimester, the potential adverse effect of SARS-CoV-2 infections in the first and second trimesters remains to be investigated.

In response to this unprecedented COVID-19 outbreak in Wuhan, confirmed and suspected COVID-19 infection has been included as one indication for cesarean section in our hospital, because there was only one negative pressure operation room suitable for airborne precautions. Two patients had vaginal delivery in labor rooms of positive pressure before they were diagnosed with COVID-19 pneumonia. No transmission events occurred in the doctors and midwives, who were wearing a full set of personal protective equipment (N95 respirators, protective gown, gloves and goggles) during the delivery procedure. Healthcare workers need to stay vigilant against COVID-19 infection when there is an epidemic in neighborhood or pregnant women have a travel history to an epidemic area within 14 days.

As suggested by Favre *et al*,[19] vaginal delivery could be considered for the benefit of patients, when there is a labor room properly equipped for airborne precautions. All healthcare workers in close contacts should strictly adhere to contact and airborne precautions in addition to standard precautions.

Similar to two previous reports of nine and one pregnant women with confirmed COVID-19 infection [7, 20], we did not find any evidence to support the vertical transmission of SARS-CoV-2 from mother to fetus via placenta or during cesarean section. Our study also added some evidence to suggest that the risk of vertical transmission during vaginal delivery might also be trivial. There were two patients with vaginal delivery, one was two days before the onset of symptoms and another was during the course of illness. Neither of their newborns had respiratory systems after birth. Unfortunately, none of them gave us the consent to collect the respiratory specimens of their neonates. Given the small sample size of our study, the possibility of vertical transmission during vaginal delivery still cannot be ruled out.

There are a few caveats in our study. First, this is a retrospective case control study from one single center, which could be subject to recall bias and selection bias. Second, we collected the data of sixteen pregnant women with laboratory confirmed COVID-19 pneumonia and eighteen suspected cases with typical CT imaging. Although this is the largest number of pregnant women with COVID-19 pneumonia in literature so far, the sample size is still relatively small. Nevertheless, given the potential of this virus to cause a global pandemic, we believe our study could be one of important clinical studies to guide the clinical diagnosis and treatment to this vulnerable group.

## Conclusion

In this study, we did not find any evidence to suggest that COVID-19 pneumonia causes severe maternal and neonatal complications among pregnant women who had vaginal delivery or caesarean section. Few patients presented respiratory symptoms on admission. The profile of laboratory investigations was not different from pregnant women without pneumonia, except a transient increase of WBC, neutrophils and CRP was found in postpartum blood tests. Given the time delay in PCR tests, chest CT scans in the third trimester might be an effective way of screening for COVID-19 pneumonia in pregnant women, particularly in areas with ongoing epidemics.

## Data Availability

All data and materials used in this work were available based on request.

## Authors’ Contributions

LYang and MP originated and designed the study. YLv, KL LYue, QL and YO contributed to data collection and clean. LH conducted data analysis. NL, LH, GS, LC and LYang interpreted the findings and drafted the manuscript. All the authors proved the final version of this manuscript.

## Ethical consideration

The ethical approval has been obtained from the ethics committee of the Hubei Provincial Maternal and Child Health Center.

## Data sharing

All data and materials used in this work were available based on request.

